# Profiling the littlest movers: Quantity and predictors of toddlers’ physical activity levels measured using a novel machine learning model

**DOI:** 10.64898/2026.01.30.26345134

**Authors:** Elyse Letts, Sara King-Dowling, Natascja Di Cristofaro, Patricia Tucker, John Cairney, Katherine M. Morrison, Brian W. Timmons, Joyce Obeid

## Abstract

**Objective:** The objectives of this study were to: (1) quantify toddlers’ total physical activity (TPA) and guideline adherence using a machine learning method; and (2) explore socio-ecological predictors (e.g., sex, childcare) of TPA.

**Methods:** Toddlers (n=103, 21.4 ± 6.9 months, 52% female) from the Hamilton, Canada region completed a gross motor assessment (Peabody Developmental Motor Scales 2^nd^ ed; PDMS-2) and wore an ActiGraph wGT3X-BT accelerometer on the right hip for 4-8 days. Parents completed demographics and physical activity surveys. TPA was estimated using a validated machine learning model and reported using descriptive statistics. Multiple linear regression explored potential predictors of TPA: age, sex, household income, older sibling, BMI-for-age z-score, gross motor z-score, childcare arrangement, parent physical activity, and temperature, controlling for accelerometer wear time.

**Results:** Toddlers had an average of 200.3 ± 44.0 minutes of daily TPA. Most (72%) met the PA guideline of 180 min/day when averaged across days, while only 27% met the guideline on all days. The regression model was significant and explained 57% of the variation in TPA (F_13,79_ = 8.09, *p* < 0.0001). Controlling for wear time, the only significant positive predictors were age and PDMS-2 z-score.

**Conclusion:** Almost three quarters of toddlers met the TPA guidelines. Older toddlers and toddlers with more advanced gross motor skills for their age participated in more daily TPA. Future research should continue to apply machine learning methods in more diverse samples and could build on modifiable predictors (e.g., motor skill) to design interventions to improve toddlers’ PA levels.

## Introduction

International physical activity (PA) guidelines recommend that toddlers (ages 12-35 months) engage in at least 180mins of daily total PA (TPA).^1,2^ These recommendations are made to ensure young children are moving enough to support their health, growth, and development.^3,4^ Estimates of the proportion of toddlers meeting these guidelines vary substantially. In a 2020 systematic review, 20 studies were identified that used device-based measurement to assess the PA and sedentary time (SED) of toddlers. From the meta-analysis, toddlers engaged in an average of 246mins/day of TPA (subdivided into light with 194mins/day, and moderate-to-vigorous with 60mins/day).^5^ Individual studies, however, have reported anywhere from 0%^6^ to 98%^7^ of toddlers meet this recommendation. The conflicting evidence is likely related to variability in PA measurement approaches, and perhaps more importantly, a lack of established accelerometry protocols for the toddler population.^5^

The 2020 review^5^ identified 10 unique thresholds (cut-points) used to quantify toddler activity while a 2022 review^8^ found 28 unique cut-points for ActiGraph accelerometers were used across 480 analyses of young children (0-5 years). A limitation of these methods is that they are device specific and reduce the variability of raw data to activity counts, which are then averaged over epochs (e.g., 15s), after which the cut-points are applied. Further, there is wide variability in published cut-point thresholds themselves, with one study concluding that 18% to 98% of toddlers met PA guidelines depending on cut-point applied.^7^ Due to these limitations, many^8,9^ have recommended moving away from count-based cut-points towards methods using raw data, such as machine learning (ML). We have recently developed a toddler specific ML model,^10^ which has been shown to outperform existing cut-point methods when validated against direct observation. This novel toddler ML model additionally distinguishes time spent in non-volitional movement (NVM; e.g., carried), further improving estimation of SED and PA.^10^ Having more accurate measurements of toddlers’ PA is essential to understand toddlers’ guideline adherence. Using more accurate methods also enables us to better identify meaningful predictors of their PA.

Many multilevel factors can influence toddlers’ PA engagement. The socio-ecological model of development provides a comprehensive framework for understanding how these factors influence development and physical activity behavior^11–13^ and includes individual, social, and environmental factors. While there has been considerable investigation into these factors in school age children, the literature is relatively sparse, and with mixed findings, when looking at the toddler age range, as summarized in Table 1.

**Table 1.** Summary of previous findings on factors that may influence toddlers’ device-measured physical activity.

| Socio-ecological factor | Predictor | Relationship with PA |
| --- | --- | --- |
| Individual<br>(Demographic/Biological) | Sex | Males > Females<sup>6,14–19</sup><br>No difference<sup>7,15,20–23</sup> |
|  | Age | Older > younger<sup>6,14,15,17,24</sup><br>No difference<sup>16,25</sup> |
|  | Income | High income > low income<sup>15,24</sup> |
|  | Race/Ethnicity | White > Non-White<sup>26</sup><br>No difference<sup>16,19,21,25</sup> |
|  | Weight status | Normal weight > over- and under-weight<sup>27</sup><br>No difference<sup>6,15,19,21,25</sup> |
|  | Parent education | No difference<sup>6,7,16,20,21,25</sup> |
|  | Marital status | No difference<sup>6,16,19,28</sup> |
| Individual (Behavioural<br>Attributes/Skills) | Motor skill | Improved motor skill > other<sup>17,19,29</sup><br>No difference<sup>6,14,20,21</sup> |
|  | Sleeping | Less sleep > more sleep<sup>24</sup><br>No difference<sup>17</sup> |
|  | TV/Screen time | Less TV/screen time > more TV/screen time<sup>7,28</sup><br>No difference<sup>6,14,19,22,30</sup> |
| Individual<br>(Psychological/emotional) | Activity temperament<br>(psychological component of preferred levels of activity) | Higher levels of preferred activity > lower levels of preferred activity<sup>29</sup><br>No difference<sup>6,31</sup> |
|  | Other (self-regulation, cognitive performance, emotionality) | No difference<sup>29</sup> |
| Social/Cultural | Parent PA levels | More parent PA > less parent PA<sup>21,22,28-30</sup><br>No difference<sup>31</sup> |
|  | Childcare attendance/teacher promotion of PA | Childcare attendance/teacher promotion of PA > other<sup>7,17,21,28</sup><br>No difference<sup>6,16,19,20</sup> |
|  | Time spent with peers | More time with peers > less time with peers<sup>30,32</sup> |
|  | Siblings | Having siblings (often older) > no siblings<sup>6,16,26,28</sup><br>No difference<sup>19,20</sup> |
|  | Housing type | Higher living area/person > lower living area/person<sup>17,29</sup><br>No difference<sup>28,31</sup> |
|  | Parenting practices | Support of PA > other<sup>19</sup><br>No difference<sup>14,17,22,30,31</sup> |
| Physical environment | Time in outdoor spaces | More time outside > less time outside<sup>29</sup><br>No difference<sup>30</sup> |
|  | Weather/season | Warmer weather > other<sup>21,29,33</sup><br>No difference<sup>17,34</sup> |
|  | Neighbourhood characteristics<br>(e.g., safety walkability, parks) | More play spaces > less play spaces<sup>26</sup><br>Higher perceived neighbourhood safety > lower perceived neighbourhood safety<sup>29</sup><br>No difference<sup>31,33</sup> |
|  | Population density | Decreased density > increased density<sup>35</sup> |

Clearly, there are gaps in our understanding of toddler PA and its predictors, which are likely attributable to sample or methodological differences between studies. While all studies above use accelerometry to quantify PA, they span 4 wear locations, multiple devices, and many analytic methods (e.g., different cut-points). To date, no study has investigated levels and predictors of toddlers’ PA using a ML model. The objectives of this study were to use our improved ML model to: (1) quantify toddler PA and guideline adherence in a sample of Canadian toddlers; (2) explore predictors of PA. We hypothesized that the majority of toddlers would meet the PA guidelines and that we would identify variables related to PA informed by prior research and the social-ecological model.

## Methods

### Study Overview

Toddlers were recruited as part of the cross-sectional **i**nvestigating the validity and reliability of accelerometer-based measures of **P**hysica**L A**ctivity and sedentar**Y** time in toddlers (iPLAY) study from the Hamilton, Canada area through childcare centres, community events, websites, and social media. Toddlers were 12.0 to 35.9 months of age with no known diagnosis that impacts movement. This study received ethics clearance from the Hamilton integrated Research Ethics Board (HiREB #3674). Informed written consent was obtained from a parent of each participant. Data were collected from February 2019 through March 2020.

Participants attended a study visit where height and weight were measured, a gross-motor assessment was conducted, and parents completed questionnaires. Toddlers then wore an accelerometer at home for the following 7 days.

### Outcome Measures

#### Accelerometer Data

Participants wore an ActiGraph wGT3X-BT accelerometer (dynamic range: ±8*g*; 30Hz sampling frequency) on the right hip on an elastic waist band. They were asked to wear the accelerometer for 7 days, removing it for sleep and water activities. Parents were asked to record all times the device was put on and removed in a logbook as well as nap times.

Upon accelerometer return, the raw data .gt3x files and the 1-second .agd epoched files were downloaded using ActiLife. The .agd files were then manually cleaned by comparing them to the parent logbooks to exclude times the device was not worn using a previously published method.^36^ All naps identified in the logbook were removed from the wear time. This means that the final wear time represents the awake wear time. Participants were included if they had at least 4 days of a minimum of 6 hours per day of wear.^37^

The .gt3x files and wear times were passed to the Letts Little Movers Activity Analysis tool^10^ to estimate the daily time in NVM, SED, and TPA. The tool uses a gradient boosted trees ML model to more accurately classify toddler activity. It first extracts 40 time and frequency features in 5s windows then classifies each window as NVM, SED, or TPA. Finally, it provides a daily summary of minutes of wear time, NVM, SED, and TPA. We report the average of all valid wear days in this manuscript.

Finally, we calculated the average outdoor temperature for each participant on the days that they had valid accelerometer wear. Specifically, we extracted the mean daily temperature from the Government of Canada’s Historical Climate Data website (https://climate.weather.gc.ca/) based on the Hamilton International Airport.

#### Questionnaire Information

Parents also completed a demographic/medical questionnaire and the International Physical Activity Questionnaire (IPAQ)^38^ short form. IPAQ responses were converted to categorical responses (high, moderate, or low PA) based on published scoring protocols.^39^ The demographic/medical form included age, sex, household income, and childcare type, among others. Details of how the variables used in the regression (see *Statistical Analyses* section) were classified are available in Table 2.

**Table 2.**
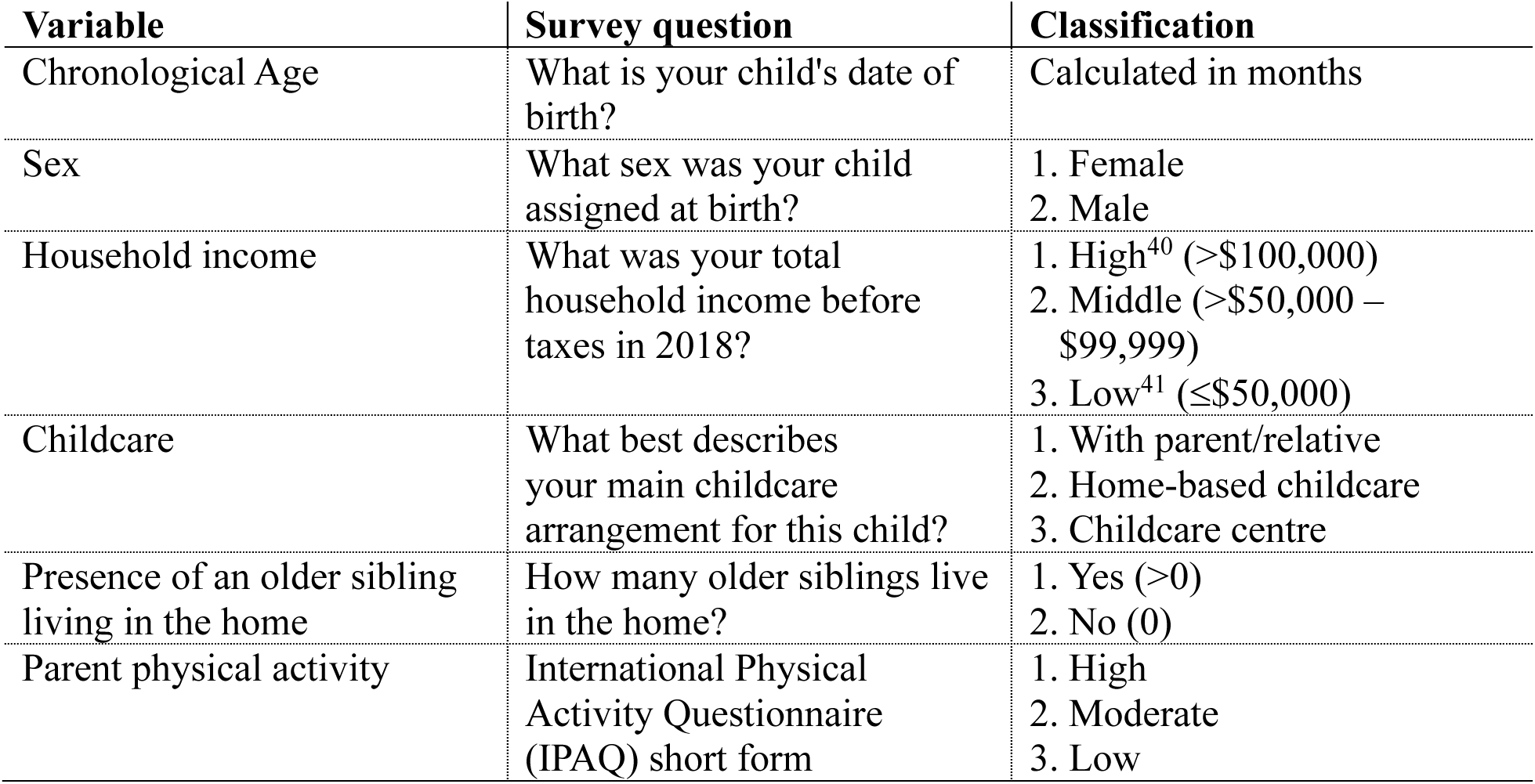
Details of questionnaire information classification.

| **Variable** | **Survey question** | **Classification** |
| --- | --- | --- |
| Chronological Age | What is your child's date of birth? | Calculated in months |
| Sex | What sex was your child assigned at birth? | 1. Female<br>2. Male |
| Household income | What was your total household income before taxes in 2018? | 1. High<sup>40</sup> (>\$100,000)<br>2. Middle (>\$50,000 – \$99,999)<br>3. Low<sup>41</sup> (≤\$50,000) |
| Childcare | What best describes your main childcare arrangement for this child? | 1. With parent/relative<br>2. Home-based childcare<br>3. Childcare centre |
| Presence of an older sibling living in the home | How many older siblings live in the home? | 1. Yes (>0)<br>2. No (0) |
| Parent physical activity | International Physical Activity Questionnaire (IPAQ) short form | 1. High<br>2. Moderate<br>3. Low |

#### Anthropometric Data

At the baseline visit, participants height/length and weight were measured. Height (to the nearest 0.1 cm; average of duplicate measures within 0.3 cm) was taken when the participant could stand independently using a portable stadiometer. Length was taken if the toddler was unable to stand independently. Weight (in kilograms) was measured either directly (able to remain still during measurement) or by being carried by the parent/researcher and subtracting the parent/researcher weight. Weight was measured at least in duplicate, or until the values were within 0.1 kg, and the average was used. The World Health Organization R package “anthro” (v1.0.1) was used to calculate body mass index (BMI) and BMI-for-age z-scores.

#### Motor Assessment

The baseline visit included a gross motor assessment, the Peabody Developmental Motor Scales – 2^nd^ ed (PDMS-2).^42^ The PDMS-2 is a validated assessment for children ages 1 month through 6 years and contains six subscales, four of which assess gross motor development. Age-standardized (adjusted for prematurity) scores from three subscales (stationary, locomotor, and object manipulation [>12 months adjusted age] or reflexes [<12 months adjusted age]) are summed and converted to age-specific z-scores from the PDMS-2 manual.^42^ All assessments were conducted by trained research staff who held undergraduate or doctoral degrees.

### Statistical analyses

For objective 1, to quantify toddler TPA and guideline adherence using the improved ML method, we used descriptive statistics to characterize the average daily TPA in minutes and the proportion of toddlers meeting the 180mins/day guideline. Based on recent literature,^43^ we considered two interpretations of the guidelines where (A) toddlers had an average^5^ of 180mins/day across all days, and (B) toddlers had 180mins of TPA on each^7^ day. For objective 2, we conducted a multiple linear regression to explore social-ecological predictors of ML-quantified average daily TPA. Predictors selected were: age (demographic/biological), sex (demographic/biological), household income (demographic/biological), BMI-for-age z-score (demographic/biological), PDMS-2 z-score (behavioural attributes/skills), childcare arrangement (social/cultural), parent PA (social/cultural), older sibling (social/cultural), and mean outdoor temperature (physical environment), controlling for accelerometer wear time.

Predictors were limited to ensure sufficient statistical power and were selected to cover all domains of the socio-ecological model (except psychological; not assessed in this study) based on previous investigations (theoretical foundations) and available information provided by families in this study. Specifically, the variables listed above contain a total of 13 parameters (continuous variable = 1 parameter; categorical variable = # of levels – 1 parameters). With 13 parameters, 101 participants are needed to detect a medium to large effect size (f^2^ = 0.2) with 80% power and α = 0.05. Given our limited sample size, it would also not be appropriate to conduct a univariate screening as this would inflate Type I error, may bias regression coefficients, and compromise the validity of *p*-values.^44^

## Results

Of the 111 participants, 109 participants had useable data (2 datafiles were corrupt), and 103 met the minimum wear time and days. Included participants had an average age of 21.4±6.9 months and 52% were female. Full demographic details are available in Table 3. Toddlers had an average of 200.3±44.0mins of daily TPA and 72% (n=74) met the PA guideline when calculated as average daily minutes of TPA (see Figure 1 Panel A). When the guideline was assessed as meeting 180mins of TPA on all valid days, only 27% (n=28) met the guidelines. Figure 1 Panel B shows the number of days participants met the guidelines relative to their valid wear days.

**Figure 1.**
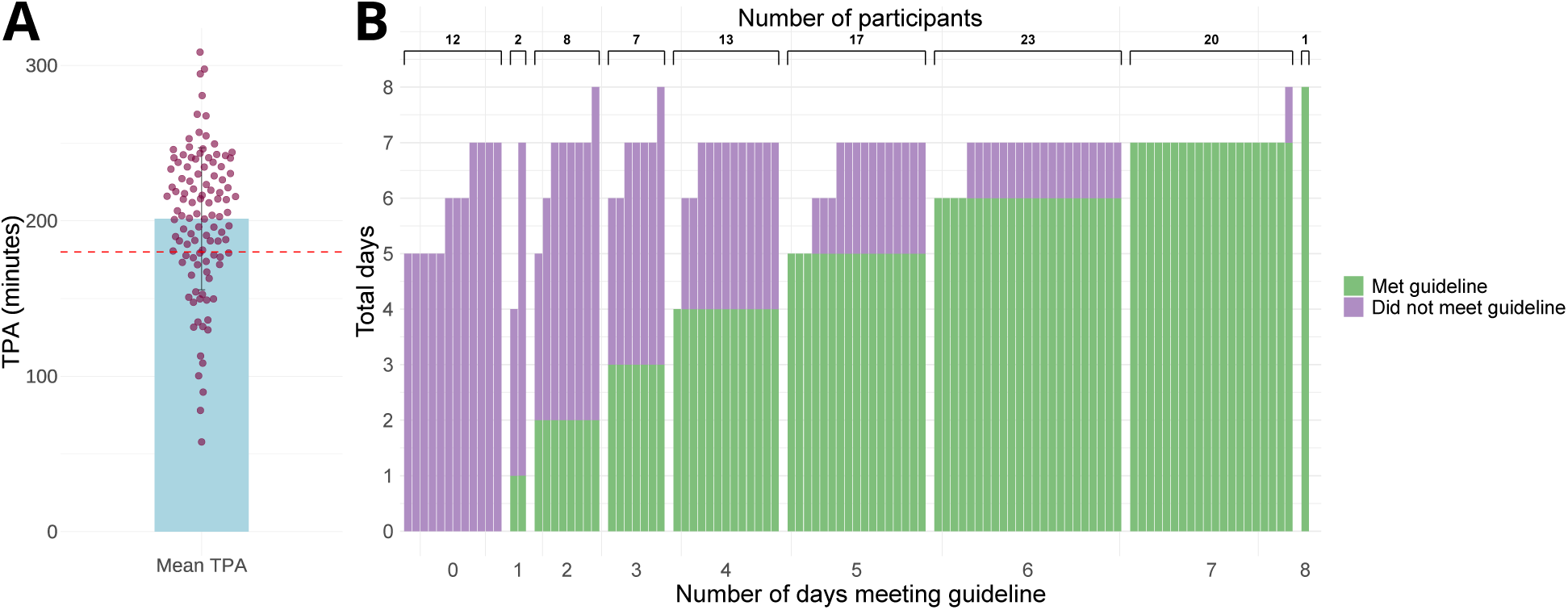
Average daily toddler total physical activity (Panel A) and participant adherence to the 180 minutes per day of TPA guideline by number of valid days (Panel B). In Panel A, the bar represents the average daily TPA across all participants, the dots represent the individual toddler’s average daily TPA, and the red line represents the 180 minutes guideline. In Panel B, participants are grouped by number of days they met the guidelines. Each bar represents one participant and shows the number of days they met the guideline (green) and the number of days they did not meet the guideline (purple).

**Table 3.** Participant characteristics.

|  | Overall<br>(N=103) |
| --- | --- |
| **CHILD DETAILS** |  |
| **Daily accelerometer wear time (min)** |  |
| Mean (SD) | 567 (56.8) |
| Median [Min, Max] | 570 [421, 712] |
| **Daily total physical activity (min)** |  |
| Mean (SD) | 200 (44.0) |
| Median [Min, Max] | 205 [57.7, 298] |
| **Average daily temperature per participant across days with valid accelerometry (°C)** |  |
| Mean (SD) | 6.9 (9.6) |
| Median [Min, Max] | 5.3 [-11.0, 23.6] |
| **Age (months)** |  |
| Mean (SD) | 21.4 (6.9) |
| Median [Min, Max] | 19.4 [12.0, 36.2] |
| **Sex [N (%)]** |  |
| Female | 53 (51.5%) |
| Male | 50 (48.5%) |
| **BMI-for-age z-score** |  |
| Mean (SD) | 0.66 (1.01) |
| Median [Min, Max] | 0.55 [-1.39, 3.54] |
| **PDMS-2 gross motor z-score** |  |
| Mean (SD) | 0.06 (0.54) |
| Median [Min, Max] | 0.13 [-1.73, 1.40] |
| **Race/Ethnicity [N (%)]** |  |
| White | 74 (71.8%) |
| Multiracial | 22 (21.4%) |
| Asian | 5 (4.9%) |
| Latin American | 2 (1.9%) |
| **Childcare [N (%)]** |  |
| With parent/relative | 68 (66.0%) |
| Childcare centre | 25 (24.3%) |
| Home-based childcare | 10 (9.7%) |
| **Older sibling [N (%)]** |  |
| No | 60 (58.3%) |
| Yes | 43 (41.7%) |
| **PARENT DETAILS** |  |
| **Household income [N (%)]** |  |
| High | 55 (53.4%) |
| Middle | 31 (30.1%) |
| Low | 15 (14.6%) |
| Not reported | 2 (1.9%) |
| **Perceived neighbourhood quality [N (%)]** |  |
| Excellent | 73 (70.9%) |
| Good | 22 (21.4%) |
| Average | 7 (6.8%) |
| Poor | 0 (0.0%) |
| Very poor | 0 (0.0%) |
| Not reported | 1 (1.0%) |
| **Parent physical activity (IPAQ) [N (%)]** |  |
| High | 46 (44.7%) |
| Moderate | 40 (38.8%) |
| Low | 10 (9.7%) |
| Not reported | 7 (6.8%) |
| **Primary language at home [N (%)]** |  |
| English | 95 (92.2%) |
| Other (Italian, Khmer, Mandarin, Portuguese, Punjabi,<br>Spanish) | 7 (6.8%) |
| Not reported | 1 (1.0%) |
| **Marital status [N (%)]** |  |
| Married | 84 (81.6%) |
| Living common-law/with partner | 15 (14.6%) |
| Single, never married | 2 (1.9%) |
| Separated | 1 (1.0%) |
| Not reported | 1 (1.0%) |
| **Parent 1 (completing survey) highest education [N (%)]** |  |
| College/university | 64 (62.1%) |
| Graduate/professional degree | 36 (35.0%) |
| High school | 2 (1.9%) |
| Not reported | 1 (1.0%) |
| **Partner/spouse of parent 1 (not completing survey) highest**<br>**education [N (%)]** |  |
| College/university | 65 (63.1%) |
| Graduate/professional degree | 28 (27.2%) |
| High school | 6 (5.8%) |
| Not reported | 4 (3.9%) |

For objective 2, the multiple linear regression met assumption checks of homoscedasticity (plot of studentized residuals versus unstandardized predicted values), normal distribution of residuals (Q-Q plot, Shapiro-Wilk *p*=0.929), no multicollinearity (variance inflation factor <5), no autocorrelation (Durbin-Watson=1.989), and all observations had a Cook’s distance less than 0.2. The overall model was significant (F_13,79_=8.09, *p*<0.001) and explained 57% (R^2^=0.571) of the variation in TPA. R^2^ changed from 0.240 with wear time alone to 0.571 including all predictors. Controlling for wear time, the only significant individual predictors were age (B=2.0, SE=0.5, *p*<0.001) and PDMS-2 gross motor z-score (B=24.5, SE=6.8, *p*<0.001). Sex, household income, BMI-for-age z-score, childcare, parent PA, presence of an older sibling, and outdoor temperature were not significant predictors. Table 4 provides the full results from the regression and Figure 2 shows the plots for each individual predictor.

**Figure 2.**
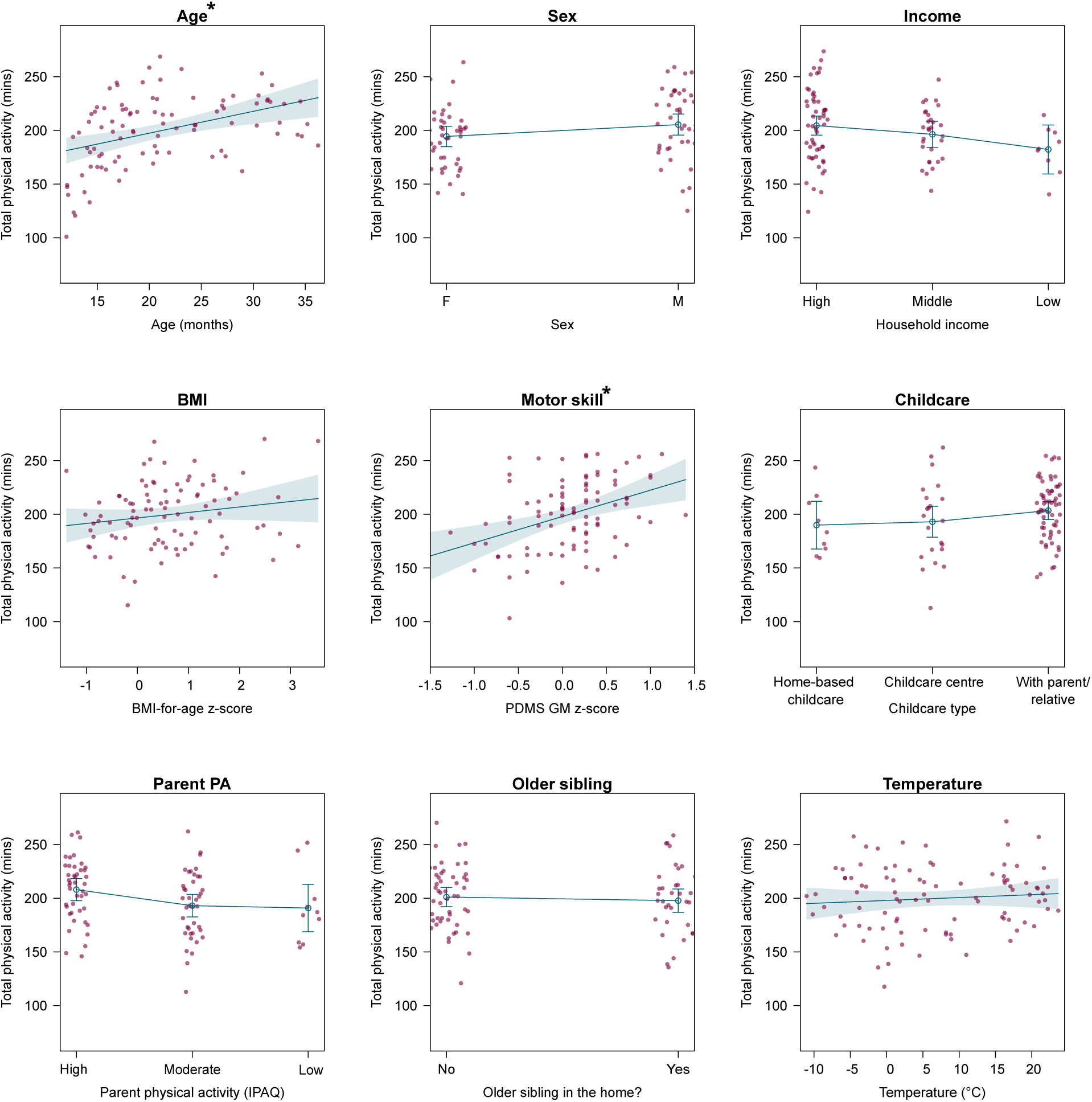
Effect plots of all predictors in the multiple linear regression model. Asterisks (*) represent significant predictors. Circles represent individual participants, and blue lines show the regression and 95% confidence intervals.

**Table 4.** Results from the multiple linear regression exploring potential predictors of toddlers’ physical activity.

| Category | Variable | B | Standard-<br>dized B | SE | t value | p-value |
| --- | --- | --- | --- | --- | --- | --- |
|  | (Intercept) | -42.4 | NA | 40.5 | -1.0 | 0.299 |
|  | Wear time | 0.3 | 0.4 | 0.1 | 5.0 | **<0.001** |
| Demographic/<br>biological | Age | 2.0 | 0.3 | 0.5 | 3.7 | **<0.001** |
|  | Sex | 11.0 | 0.1 | 7.0 | 1.6 | 0.120 |
|  | HouseholdIncome:Middle | -8.2 | -0.1 | 7.7 | -1.1 | 0.290 |
|  | HouseholdIncome:Low | -22.3 | -0.1 | 12.4 | -1.8 | 0.077 |
|  | BMI-for-age z-score | 5.1 | 0.1 | 3.6 | 1.4 | 0.162 |
| Behavioural<br>attributes/skills | PDMS-2 z-score | 24.5 | 0.3 | 6.8 | 3.6 | **0.001** |
| Social/cultural | Childcare:Childcare centre | 3.1 | 0.0 | 13.3 | 0.2 | 0.816 |
|  | Childcare:With parent/relative | 13.5 | 0.1 | 12.0 | 1.1 | 0.263 |
|  | ParentPA:Moderate | -15.0 | -0.2 | 7.6 | -2.0 | 0.053 |
|  | ParentPA:Low | -17.1 | -0.1 | 12.5 | -1.4 | 0.176 |
|  | OlderSibling | -3.4 | 0.0 | 7.3 | -0.5 | 0.644 |
| Physical<br>environment | Mean daily temperature | 0.3 | 0.1 | 0.4 | 0.7 | 0.480 |

## Discussion

The purpose of this study was to (1) quantify toddler PA and guideline adherence in a sample of Canadian toddlers using ML and (2) explore predictors of PA. In our sample of 103, toddlers had an average of 200.3±44.0mins of TPA and 72% had an average daily TPA of at least 180mins. However, only 27% had at least 180mins of TPA on all valid days. We also found that older toddlers and toddlers who had a higher gross motor z-score, independent of age, obtained more TPA.

A positive finding of this study is that nearly three quarters of our participants did meet the PA guidelines. Previous research in similar samples found that between 0%^6^ to 98%^7^of toddlers met the guideline on average. Our results compared most closely to a study that found that 76%^45^ of toddlers met guidelines using cut-point techniques. While most toddlers in our sample had an average of at least 180mins/day, about one quarter did not meet this threshold. In fact, 12% of toddlers (mean±SD (range) age: 14.6±4.0 (12.0-26.5) months) did not achieve 180mins of TPA on *any* day. While this 12% includes many of the younger toddlers (who may not be walking yet), daily movement is essential for toddlers’ health and development of motor skills.^3,4^ PA levels in the early years track into childhood^46^ and even into adulthood^47^, meaning these children may be at risk for suboptimal development and poor health outcomes.

When looking at predictors, our findings support the existing research in that age^6,14,15,18,22,24,29^ and motor skills^17,19,29^ are associated with amount of daily TPA. Most toddler studies find higher PA with increasing age (even when accounting for awake wear time differences^6,22,24,29^), likely tied to skill acquisition; as toddlers learn to walk, run and jump, their movements get bigger and faster, increasing their number of activities considered TPA. Toddlers also require less sleep as they age, meaning they may have more time to engage in PA. This also aligns with our motor skill findings: at any given age, those with a higher gross motor z-score are likely to accumulate more TPA. This makes sense as the ML model captures a wide range of activities considered TPA such as crawling, walking, squatting/standing, and stair climbing.^10^ There is also likely a bi-directional effect of motor skill and PA. Toddlers who accumulate more PA have more opportunity to practice new skills leading to improved gross motor skill acquisition^3^ and those with improved gross motor skill have likely acquired more physically active skills (e.g., running). Opposingly, not all studies in toddlers have found a relationship between motor skill and PA.^6,14,20,21^ However, these studies all use a dichotomized variable of high/low motor skill or milestone attained/not attained (across both standardized in-person assessments and parent report), while studies that do a see a difference tend to use a continuous variable, as was done in this study.^17,29^ Ultimately, these findings suggest that future interventions should support both toddlers’ PA and motor development.

We found no significant independent impact of sex on PA levels, inconsistent with much of the literature.^6,14–19^ While not statistically significant, in our study, boys accumulated an average of 14 more minutes per day of TPA than girls (mean±SD: 207.9±46.7 vs. 193.2±40.4mins). We know that sex/gender differences in PA exist in school-age children and even in children as young as preschoolers,^12,48,49^ but findings in toddlers have been more mixed. Some studies in toddlers have found that boys^6,14–19^ are more active, while others find no difference.^7,15,20–23^ Our data support that the toddler years may be an emerging time for these sex-based differences, while the difference is not yet significant. This means that the toddler and preschool years may be a critical time^46^ to intervene to minimize sex-based PA discrepancies.

Similarly, we found no difference in TPA by child BMI, childcare arrangement, household income, older siblings, nor parental PA. This reinforces existing literature that in toddlers, BMI does not appear to influence PA.^6,15,18–23,25^ That being said, our sample was mostly considered normal weight, with only 17% above the 95^th^ percentile for BMI-for-age. As such, results should be interpreted with caution and future work should investigate the TPA of toddlers living with diagnosed obesity. For childcare arrangement, previous studies have shown that some specific attributes within a childcare centre are more likely to influence PA including quality of the outdoor environment, designated PA time, and site-specific pedagogy.^32^ The level of detail we had available did not distinguish these details. There may also be an influence of age and childcare arrangement that could obscure this relationship, whereby older children are more likely to be in childcare. This should be investigated in future work. We observed no significant difference in TPA between toddlers from low (193.2±40.3mins), middle (197.1±49.0mins), and high-income (204.7±43.0mins) families. It should be noted that the sample was mostly middle and high income (83%) so these results should be interpreted with caution. When looking at parent PA as measured by the IPAQ, we saw no statistically significant differences between parents with low (164.1±64.7mins), moderate (193.0±42.8 mins), and high (212.6±36.5 mins) levels of activity. However, this does represent a difference of 48mins/day of toddler PA between parents with low compared to high activity. As parent PA was measured with a survey in this analysis (and only from one parent), device-based measures of parent PA should be used to further explore this relationship.

Finally, looking at the environment, we included outdoor temperature based on a prior study^33^ that found that temperature was the only environmental variable significantly related to toddler PA. Our study opposes these findings as we saw no significant impact of temperature. The study that did find an impact of temperature reported a lower mean temperature (−0.33±9.83°C) than in our study (6.9±9.6°C) which may have resulted in more indoor time to avoid the colder temperatures. We also did not assess other environmental factors such as walkability score, nearby parks, and neighbourhood safety,^29,33^ which should be investigated in future work.

### Limitations

There are some key methodological issues to consider when interpreting our findings. First, while the ML model used has been shown to improve accuracy of toddler PA measurement, in this sample, it has not yet been validated in an independent sample. Second, our sample was mostly White, and from English-speaking, two-parent, and highly educated households which limits the generalizability of our findings. Third, given our sample size, we could not test all potential predictors (although we were able to test nine with prior evidence). Future research should apply ML methods in more diverse samples to further explore toddlers’ PA.

### Clinical and research implications

This study applies a toddler-specific ML model to provide more accurate estimates of toddlers’ PA guideline adherence in a sample of 103 toddlers. This fills an important gap in our understanding of toddlers’ activity levels and suggests that most young children are meeting current PA guidelines. It further provides insight into predictors of toddlers’ activity, which could support the design of future clinical interventions.

## Conclusion

Using ML to quantify toddlers’ PA and explore its predictors, we found that 72% meet activity guidelines when averaged across all days, with toddlers who are younger and with a lower gross motor z-score at greatest risk for low PA. Future research should continue to investigate these findings in larger and more diverse samples and can build on modifiable predictors (e.g., motor skill) to support the design of interventions to improve toddlers’ PA levels.

## Data Availability

The datasets used and/or analysed during the current study are available from the corresponding author on reasonable request.

## Declarations

### Ethics approval and consent to participate

This study received ethics clearance from the Hamilton integrated Research Ethics Board (HiREB #3674). Informed written consent was given by a parent of each participant.

### Authors’ contributions

EL was involved in the conceptualization of this paper, analysed all data, and wrote the original draft of this manuscript. SKD was involved in the project conceptualization, methodology development, project administration, and manuscript editing. ND was involved in the project conceptualization, methodology development, project administration, data collection, and manuscript editing. PT was involved in the project conceptualization, methodology development, funding acquisition, and manuscript editing. JC was involved in the project conceptualization, methodology development, funding acquisition, and manuscript editing. KMM was involved in the conceptualization of this paper, data analysis, and manuscript editing. BWT was involved in the project conceptualization, methodology development, project administration, funding acquisition, supervision, and manuscript editing. JO was involved in the project conceptualization, methodology development, project administration, data analysis, funding acquisition, supervision, and manuscript editing. All authors reviewed and approved this version of the manuscript for submission.

## Acknowledgements

We would like to thank the participants and their families who made this research possible. We also acknowledge the efforts of Sarah da Silva and Camilla Wegrzynowska who contributed to the manual cleaning of accelerometer data.

